# Use of machine learning for comparing disease risk scores and propensity scores under complex confounding and large sample size scenarios: a simulation study

**DOI:** 10.1101/2022.02.03.22270151

**Authors:** Yuchen Guo, Victoria Y Strauss, Daniel Prieto-Alhambra, Sara Khalid

## Abstract

**Background:** The surge of treatments for COVID-19 in the ongoing pandemic presents an exemplar scenario with low prevalence of a given treatment and high outcome risk. Motivated by that, we conducted a simulation study for treatment effect estimation in such scenarios. We compared the performance of two methods for addressing confounding during the process of estimating treatment effects, namely disease risk scores (DRS) and propensity scores (PS) using different machine learning algorithms.

**Methods:** Monte Carlo simulated data with 25 different scenarios of treatment prevalence, outcome risk, data complexity, and sample size were created. PS and DRS matching with 1: 1 ratio were applied with logistic regression with least absolute shrinkage and selection operator (LASSO) regularization, multilayer perceptron (MLP), and eXtreme Gradient Boosting (XgBoost). Estimation performance was evaluated using relative bias and corresponding confidence intervals.

**Results:** Bias in treatment effect estimation increased with decreasing treatment prevalence regardless of matching method. DRS resulted in lower bias compared to PS when treatment prevalence was less than 10%, under strong confounding and nonlinear nonadditive data setting. However, DRS did not outperform PS under linear data setting and small sample size, even when the treatment prevalence was less than 10%. PS had a comparable or lower bias to DRS when treatment prevalence was common or high (10% - 50%). All three machine learning methods had similar performance, with LASSO and XgBoost yielding the lowest bias in some scenarios. Decreasing sample size or adding nonlinearity and non-additivity in data improved the performance of both PS and DRS.

**Conclusions:** Under strong confounding with large sample size DRS reduced bias compared to PS in scenarios with low treatment prevalence (less than 10%), whilst PS was preferable for the study of treatments with prevalence greater than 10%, regardless of the outcome prevalence.

**Key Messages:**

- When handling nonlinear nonadditive data with strong confounding, DRS estimated by machine learning methods outperforms PS in scenarios with low treatment prevalence (less than 10%).
- However, if having linear data and small sample size data with strong confounding, we did not observe DRS outperformed PS even when treatment prevalence was less than 10%.
- Our results suggested that using PS performed better compared to DRS in tackling strong confounding problems with treatment prevalence greater than 10%.
- Small sample size increased bias for both DRS and PS methods, and it affected DRS more than PS.

## 1 INTRODUCTION

Given the increasing abundance of routinely collected data from clinical practice, the use of observational data for understanding associations between exposures (hereafter treatments) and outcomes is on the rise [1]. However, one of the key challenges associated with observational data is the presence of known and unknown sources of confounding [2]. Propensity score (PS) analysis is one of the most popular approaches for mitigating confounding effects, where the PS for a given individual represents the probability of receiving a treatment of interest conditional on known confounders. Several PS methods have been tested to reduce confounding effects such as matching, stratification, and weighting [2] [3].

Disease risk score (DRS) analysis presents an alternative approach, where the DRS for a given individual is the predicted probability of the outcome conditional on the confounders. Let *T* denote treatment, *Y* denote outcome, and *X* denotes confounders, PS can be expressed as *P* (*T* = 1 |*X*) while DRS is *P* (*Y* = 1 |*T* = 0, *X*). PS is a function treatment whereas DRS is independent of treatment, this property motivated us to test its performance against PS under different treatment prevalences. While PS is well known and widely applied, DRS is relatively less widely applied in epidemiological literature. As such DRS could be potentially advantageous in scenarios with low treatment prevalence, where PS has been shown to underperform [4].

Although PS and DRS have been compared in specific scenarios with multiple confounders [5], little is known overall and the comparative performance of the two approaches - particular in cases of strong confounding and rare treatment prevalence - remains unclear [5] [6] [7] [8].

Further, while recent research has investigated machine learning methods for estimation of PS [9] [10] [11] [12] [13] [14] [15], the calculation of DRS has exclusively been performed using logistic regression [5]. According to some studies [9] [10] [14] [15], machine learning may be better suited to scenarios that involve large sample sizes and high data complexity. However, to our knowledge, a comparative performance of machine learning methods under such scenarios for PS and DRS has not been done.

Therefore we aimed to assess the performance of different machine learning methods for PS and DRS estimation under a range of common real-world scenarios, namely different treatment prevalence and different outcome risk scenarios in the presence of strong confounding.

The main contributions of this paper are to demonstrate:

- the comparative performance of PS and DRS in case of different treatment prevalence especially low treatment prevalence and under strong confounding,
- the effects of data complexity and sample size on the accuracy of PS and DRS in estimating treatment effects, and
- the comparative performance of different machine learning methods in PS and DRS estimation.

## 2 METHODS

A Monte Carlo simulation study was conducted. The simulation settings of nonlinearity and nonadditivity in [15] were followed and adapted for the various scenarios under investigation.

### 2.1 Data Simulation

We simulated datasets using two different strategies: 1. independent covariates without nonlinearity and nonadditivity, shown in the figure on left of Figure 1. 2. Covariates with nonlinearity and nonadditivity, shown in the figure on right of Figure 1. Specifically, for the second simulation strategy, we included 15 (30% of the total amount of confounders) two-way interactions for the nonadditive terms and 5 cubic terms for the nonlinear terms. For both linear and nonlinear data settings, the dataset included 50 confounders that affected both treatment and outcome, 5 instrumental variables that affected only treatment, and 1 risk factor that only affected the outcome. Details of simulation settings are shown in Table 1 and in the Appendix.

**Table 1.** Simulation settings

|  |  |
| --- | --- |
| Standard normal confounders | $X_1, \dots, X_{25}$ |
| Binomial confounders | $X_{26}, \dots, X_{50}$ |
| nonadditivity terms<br>between normal distribution<br>and binomial distribution | $X_6 * X_{26}, X_7 * X_{27}, \dots, X_{10} * X_{30}$ |
| nonadditivity terms<br>between normal distribution<br>and binomial distribution | $X_{11} * X_{12}, X_{13} * X_{14}, \dots, X_{19} * X_{20}$ |
| Nonlinear terms:<br>cubic terms | $X_1^3, X_2^3, \dots, X_5^3$ |
| nonadditivity terms<br>between cubic terms and<br>binomial distribution | $X_1^3 * X_{31}, X_4^3 * X_{34}, \dots, X_5^3 * X_{35}$ |
| Standard normal instrumental variables | $X_{66}, \dots, X_{70}$ |
| Binomial risk factor | $X_{71}$ |

**Table 2.** Simulation scenarios

| Scenarios # | Treatment prevalence | Number of Observations | Data distribution | Outcome prevalence |
| --- | --- | --- | --- | --- |
| 1 – 5 | 0.01, 0.05, 0.1, 0.25, 0.5 | 500 | Linear | 50% |
| 6-10 | 0.01, 0.05, 0.1, 0.25, 0.5 | 10000 | Linear | 50% |
| 11-15 | 0.01, 0.05, 0.1, 0.25, 0.5 | 500 | Nonlinear | 50% |
| 16-20 | 0.01, 0.05, 0.1, 0.25, 0.5 | 10000 | Nonlinear | 50% |
| 21-25 | 0.01, 0.05, 0.1, 0.25, 0.5 | 10000 | Nonlinear | 2% |

**Fig. 1.**
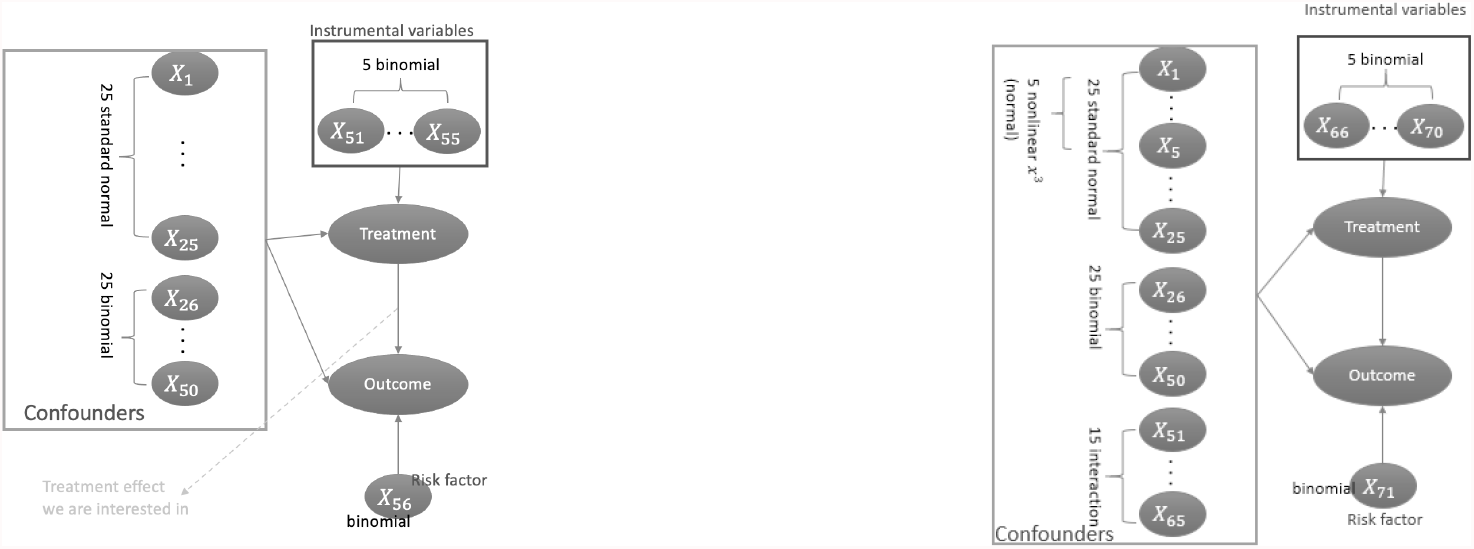
Linear and nonlinear data simulation demonstration

**Fig. 2.**
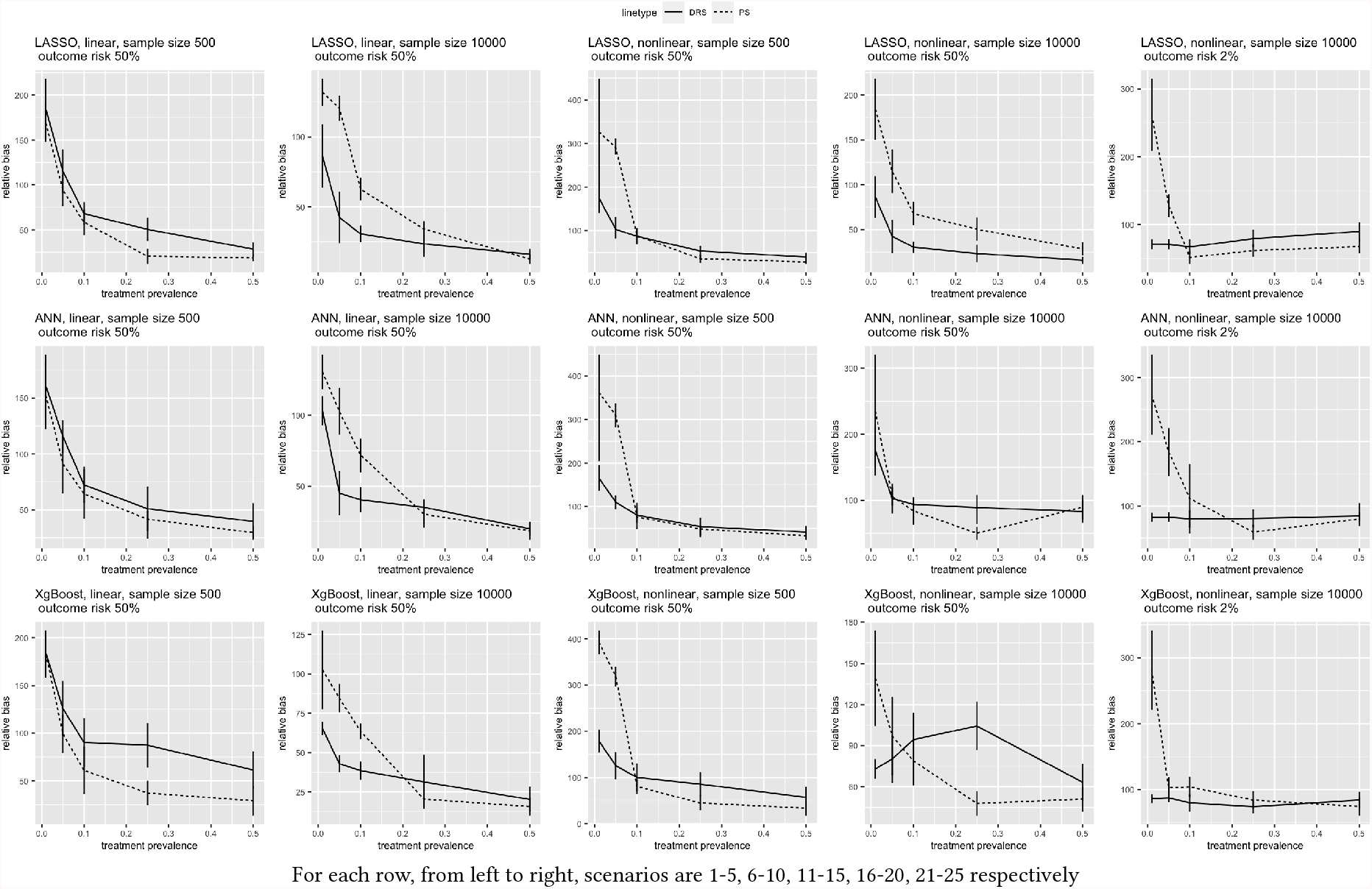
Comparison between PS and DRS under different scenarios

In total, 25 different scenarios were tested, as shown in Table 1. Different treatment prevalence ratios, outcome risk percentages, sample size, data complexities were examined. These scenarios were investigated using different settings (Table 1) including “strong confounding” and “large sample size” settings, defined as follows. Previous work typically included 10 confounders, so here we included 50 linear and non-linearly distributed confounders to simulate a “strong confounding” setting. Further, when testing large sample sizes, previous work [16] [17] used 10000 samples despite having fewer confounders than our settings, so here we used a sample size of 10000 to test a “large sample size” setting in addition to a sample size of 500.

### 2.2 Propensity Score Settings

PS represents the probability of receiving treatment of interest conditional on confounders, and many PS methods have been tested to reduce confounding effects such as matching, stratification, and weighting [2] [3]. Traditionally, PS is calculated using logistic regression with pre-selected confounders. In this study, we used machine learning methods LASSO, XgBoost, and MLP to estimate PS. Target variable was set to treatment and all 50 confounders, 5 instrumental variables and 1 risk factor were input into machine learning model. After obtaing PS, a matched cohort was obtained with 1:1 matching ratio with greedy nearest neighbour matching and without replacement.

### 2.3 Disease Risk Score Settings

DRS was first proposed by Miettinen in 1976 [18] and it is the probability of getting potential outcome conditional on confounders if untreated. It can be computed in two different ways [5]: from the unexposed population or the full cohort. Full cohort DRS is obtained by regressing outcome to covariates and treatments using the entire study population *Y* ∼ *X, T*, and then fitted values *P* (*Y* = 1 | *T* = 0, *X*) of full population are computed by setting treatment status to unexposed *T* = 0. Unexposed DRS is computed by firstly regressing outcome to covariates *Y* ∼ *X* only for unexposed population, which was done using machine learning methods as described above. Then fit the model to the entire population, fitted values *P* (*Y* = 1 | *X*)are the unexposed DRS. Same as PS, 1:1 matching was used with greedy nearest neighbour matching and without replacement.

Compared to unexposed DRS, full cohort DRS has shown better performance in terms of bias when estimating treatment effect [5], therefore it was used in this work.

### 2.4 Machine Learning Methods

To estimate PS and DRS, we applied three different machine learning methods namely least absolute shrinkage and selection operator (LASSO) regularization, multilayer perceptron (MLP), and eXtreme Gradient Boosting (XgBoost). We input all variables in the simulated dataset into our machine learning models. The target variable for estimating PS was set as treatment while the target variable for estimating DRS was set as the outcome. All machine learning methods were hyperparameter tuned using 10-fold cross validation (see Appendix for details). Matching was used to obtain the estimated treatment effect.

### 2.5 Estimands and Metrics

In either PS or DRS, a matched cohort was obtained with 1:1 matching ratio with greedy nearest neighbour matching and without replacement. The estimated treatment effect was calculated in the matched cohort via a logistic regression with only treatment as the predictor variable. The true treatment effect was calculated via a logistic regression with treatment, 50 confounders and one risk factor as the predictor variables. To evaluate the treatment effect estimation, we used relative bias, which was defined as:

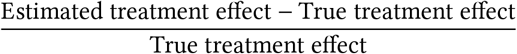

We also used the corresponding 95% confidence interval to evaluate the results.

## 3 RESULTS

### Effect of Treatment Prevalence

Comparison between PS and DRS regarding treatment prevalence are shown in Figure 2. The relative bias for PS increased with decreasing treatment prevalence, regardless of the machine learning methods used. Secondly, DRS increasingly outperformed PS with the decreasing treatment prevalence. We observed that the treatment effect estimated by DRS had a much lower bias compared to PS when having nonlinear data and treatment prevalence was lower than 10%, for all three machine learning methods. For example, DRS estimated by LASSO yielded 42.43% bias whereas PS estimated by LASSO resulted in 120.71% bias, under scenarios 6-10, when treatment prevalence was 5%. This was not true in the case of linear data, leftmost plot in Figure 2. Here PS consistently outperformed DRS, although the performance of DRS was similar to PS with the decrease of treatment prevalence. We also found that when outcome risk was rare (2%) or high(50%) and treatment prevalence was between common and high (10% - 50%), there was at least one machine learning method estimated PS can achieve lower or comparable bias compared to DRS. For instance, referring to Figure 2, scenarios 11-15 when treatment prevalence was between 10% and 50%, all PS results (dashed lines) were lower than DRS in solid line. Scenarios 16-20 when treatment prevalence was between 10% and 50%, PS estimated by method XgBoost and MLP were achieving lower bias in the dashed line below the DRS (solid lines).

In general, results show that DRS was resulted in less bias in estimation of treatment effect when both treatment prevalence and outcome risk were relatively rare, whereas PS performed better (i.e. resulted in less bias) when treatment prevalence was common, under our setting of strong confounding and nonlinear nonadditive data.

### Effect of Data Complexity

Regarding the effect of nonlinearity and nonadditivity on estimation results, Figure 4 compares how PS and DRS were affected by the nonlinearity and nonadditivity. For all methods, the bias generated by nonlinear data using PS was higher than the linear one. The difference was even more significant when the treatment prevalence was lower than 10%. For example, when treatment prevalence was 1%, the relative bias for nonlinear PS was 326.98% while the linear data generated 168.98%. Similarly, for DRS, nonlinear data introduced more bias into DRS results especially when treatment prevalence was lower than 10%.

### Effect of Sample Size

Considering how sample size affected treatment effect estimation bias, Figure 3, when decreasing sample size from 10000 to 500, treatment effect estimation bias increased regardless of the machine learning method. However, the effect on DRS was more significant than PS, when treatment prevalence was 1%: using XgBoost, LASSO, and MLP, reducing sample size from 10000 to 500 increased bias of DRS respectively from 65.35% to 184.03%, 86.36% to 184.26%, and 103.38% to 160.59%.

**Fig. 3.**
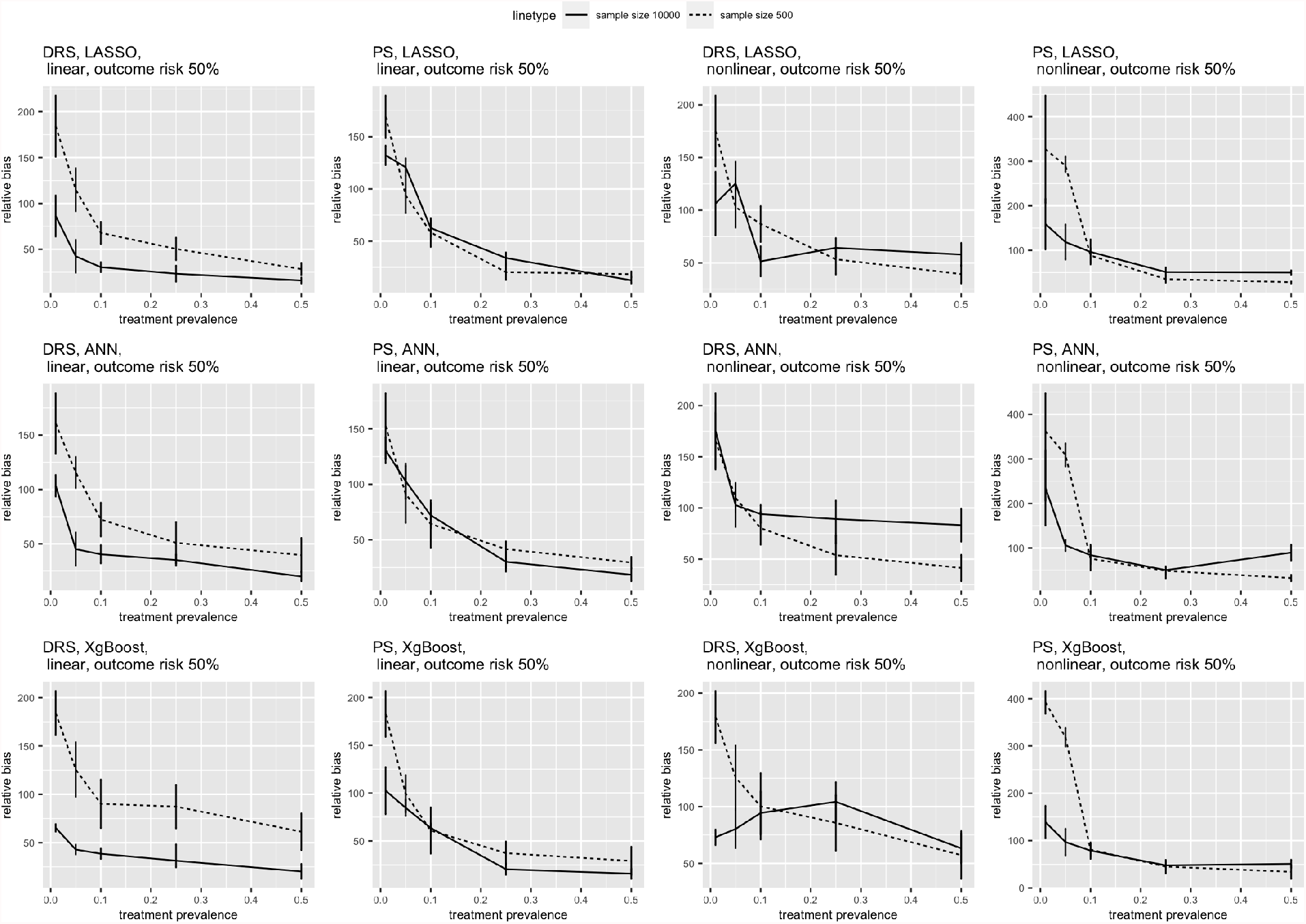
Comparison between two sample sizes: 500 and 10000

**Fig. 4.**
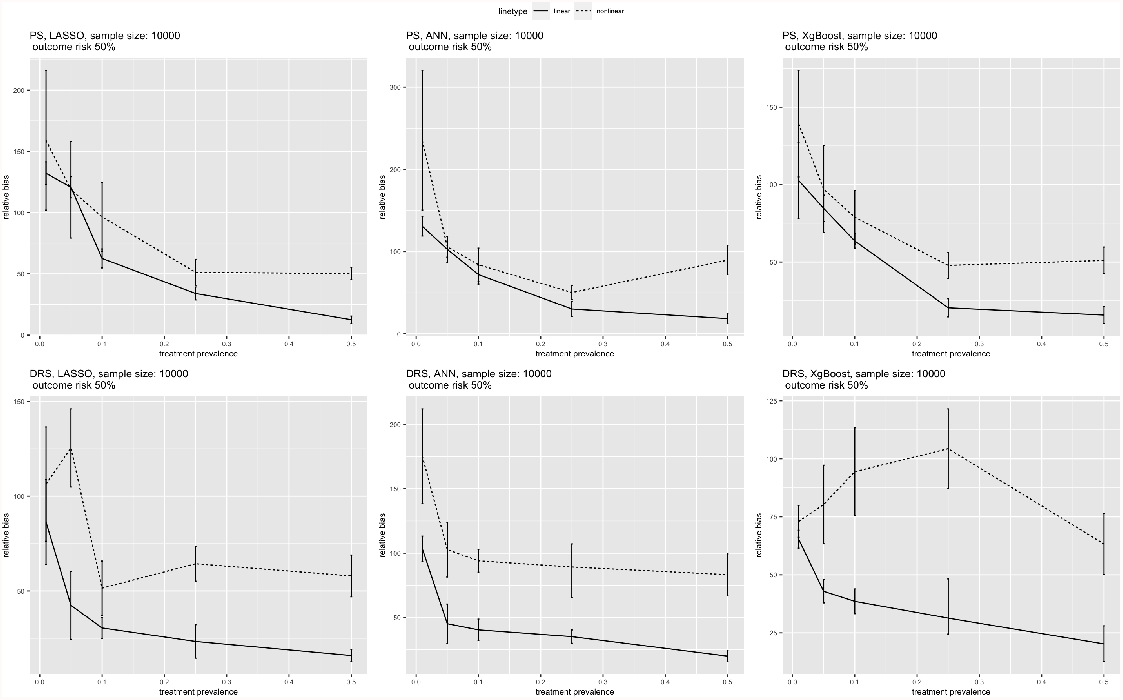
Comparison between nonlinear data and linear data

### Comparison of Machine Learning Methods

Three machine learning methods were compared too, bias generated by machine learning estimated PS and DRS are shown in Figure 5. For all the scenarios tested, three machine learning methods had similar performances, especially considering the confidence interval overlapped. However, our data suggested that there were scenarios that either XgBoost or MLP can generate the lowest bias. We observed XgBoost was giving the lowest bias when decreasing treatment prevalence lower than 10% compared to other machine learning methods, under nonlinear data with the large sample size 10000 and 50% outcome risk: At 1% treatment prevalence, PS and DRS estimated by XgBoost were giving 139.23% and 72.98% bias respectively, whereas PS and DRS estimated by LASSO were giving 158.98% and 106.36% or estimated by MLP were giving 235.23% and 175.31%; At 5% treatment prevalence, PS and DRS estimated by XgBoost were giving 97.05% and 80.31% bias respectively whereas PS and DRS estimated by LASSO were giving 118.71% and 125.43% or estimated by MLP were giving 105.96% and 102.76%. We also observed that LASSO generated the lowest bias, under 10%-50% treatment prevalence, linear data with the small sample size 500, 50% outcome risk, for both PS and DRS. For instance, referring to scenarios 1-5, at 10% treatment prevalence, PS and DRS estimated by LASSO were generating 58.35% and 67.95% relative bias respectively, whereas PS and DRS generated by XgBoost were 90.23% and 60.94% and generated by MLP were 64.31% and 72.30%. For other case scenarios, we did not observe a significant advantage of XgBoost over LASSO or vice versa with overlapping confidence intervals.

**Fig. 5.**
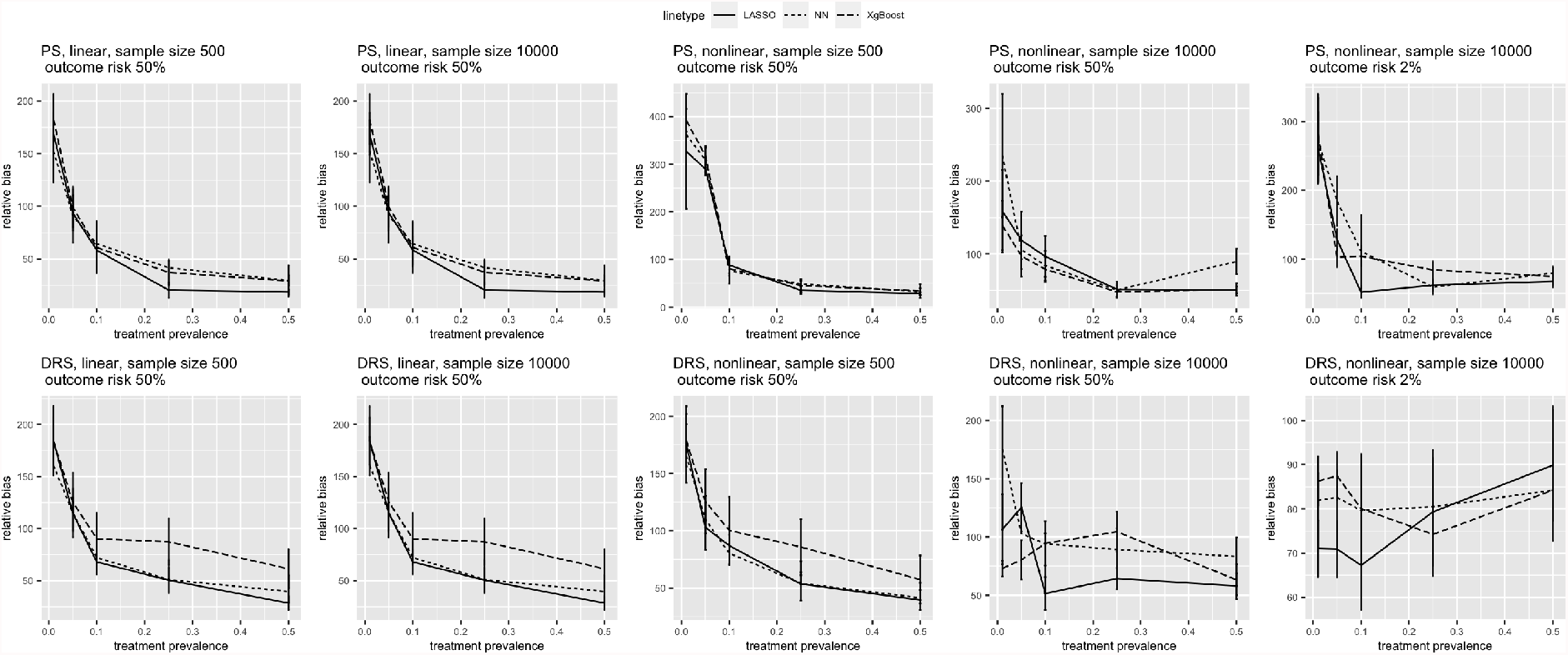
Comparison of different machine learning methods: LASSO, MLP, and XgBoost

## 4 DISCUSSION

We conducted a simulation study to 1) evaluate the performance of DRS and PS for measured confounders adjustment; 2) to assess the performance of three different machine learning algorithms in the generation of PS or DRS.

For nonlinear data under strong confounding settings, we observed that when outcome risk was high (50%) or relatively rare (2%), DRS outperformed PS when treatment prevalence was relatively rare (lower than 10%). This finding was as could be expected since models used for PS estimation may fail to capture nonlinearity and nonadditivity with the target variable (treatment prevalence) being imbalanced. We also found that when outcome risk was relatively rare (2%) or high (50%) and treatment prevalence was between common and high (10% - 50%), PS resulted in less bias compared to DRS. Previous research by Xu et al. [6] compared PS and DRS using Type 1 error and empirical power, which are other metrics that could be used in simulation studies. Type I error represents the probability of incorrectly rejecting the true null hypothesis of no treatment effect when it’s true. Empirical power is the probability of not making a Type II error, which is not rejecting the null hypothesis when it’s false. Our result was consistent with the results demonstrated by Xu et al., although that study compared PS and DRS when the treatment was common and the outcome was moderately rare or common. They found that for 1:1 matching PS outperformed DRS, as PS had lower type I error. However, their study did not have scenarios where DRS can be advantageous over PS e.g. when both treatment prevalence (lower than 10%) and outcome risk (2%) were relatively rare, as shown by our results. This could be caused by the difference in our simulation settings as well as the evaluation metric used.

Sample size also affected our estimation bias. With only 500 observations, 50 confounders, and nonlinear data setting, our data suggested that PS resulted larger bias when outcome risk was 2% and treatment prevalence was lower than 10%. This can be explained by the number of events per variable being too low, as we had 50 confounders, and when treatment prevalence was lower than 10%, we had less than one event per confounder. As some previous research demonstrated, 10 events per variable may not even be enough [19]. A previous research comparing DRS and PS [5] also demonstrated that for logistic regression, when reducing the number of events per covariate, bias increased for both PS and DRS. We also observed that sample size affected DRS more than PS. This could be expected as modelling the DRS introduced additional complexity as it requires accurately modeling the relation between treatment and outcome. Therefore, decreasing sample size could make it more difficult for DRS than PS.

Among the three machine learning methods we tested, LASSO, XgBoost, and MLP, all of them had similar performance when we had complicated nonlinear data settings. Previous review [10] found that boosting method was the most promising machine learning method for PS analysis, which was consistent with our observation that XgBoost as a boosting method outperformed the other two machine learning methods under many scenarios. We observed XgBoost as the best model when having nonlinear data, large sample size, and low treatment prevalence. We also found that LASSO was the best model when having linear data, small sample size, and common treatment prevalence. In all other cases, our data supported LASSO and XgBoost or ensemble these two methods in super learner [20]. High dimensional propensity score [13] is also a popular method to estimate PS. It includes a process of ranking covariates by their prevalence and univariate association with the outcome and/or the treatment, and it requires a specific pre-selected number of covariates. However, our approach was to allow machine learning to self-select among potentially large numbers of confounders. Additional machine learning methods such as outcome adaptive LASSO [21] and highly adaptive LASSO [22] have also been illustrated to have competitive performance compared with other popular machine learning techniques like gradient boosted machine [22], however, they were mostly compared on small scale data.

## 5 CONCLUSIONS

We compared the performance of less-known DRS with widely-used PS for estimating treatment effect in the presence of strong confounding and large sample sizes. We computed both PS and DRS using three well-established machine learning methods. These data suggest that DRS could be a preferable method compared to PS when treatment prevalence was lower than 10%, under strong confounding and nonlinear data scenarios. PS had competitive performance compared to DRS when treatment prevalence was between common and high (10% - 50%). Sample size was important when having strong confounding in data, where small sample size may lead to inaccurate estimation for both DRS and PS, and it affected DRS more than PS. Nonlinearity and nonadditivity in the data introduced more bias in both PS and DRS.

## Data Availability

Data simulation steps are included in the manuscript. All data produced in the present study are available upon request to the authors

## ACKNOWLEDGMENTS

The research was supported by the National Institute for Health Research (NIHR) Oxford Biomedical Research Centre (BRC). DPA is funded through a NIHR Senior Research Fellowship (Grant number SRF-2018-11-ST2-004). The views expressed in this publication are those of the author(s) and not necessarily those of the NHS, the National Institute for Health Research or the department of Health.

Prof. Prieto-Alhambra’s research group has received grant support from Amgen, Chesi-Taylor, Novartis, and UCB Biopharma. His department has received advisory or consultancy fees from Amgen, Astellas, AstraZeneca, Johnson, and Johnson, and UCB Biopharma and fees for speaker services from Amgen and UCB Biopharma. Janssen, on behalf of IMI-funded EHDEN and EMIF consortiums, and Synapse Management Partners have supported training programmes organised by DPA’s department and open for external participants organized by his department outside submitted work.

## Statements of ethical approval

No patient data or real data was used. This work simulated synthetic data therefore was considered exempt from IRB approval. Informed consent was not necessary at any site.

## Author contributions

YG, SK, VS, and DAP were responsible for conceiving the study. YG conducted the study analysis. All the co-authors contributed to interpreting study results and writing the manuscript. The corresponding author confirms that all authors read and approved the final manuscript

## APPENDIX

### Hyperparamters

Hyperparameters tuned for LASSO was shrinkage coefficient

Hyperparamters tuned for XgBoost were: learning rate, minimum split loss, maximum depth of a tree, minimum sum of instance weight (hessian) needed in a child, subsample ratio of the training instances

Hyperparamters tuned for MLP were: number of layers, number of units per layer, activation function, optimizer.

#### Simulation details

#### Linear data

1. Simulated independently 50 confounders from normal and binomial distributions *X*_1_, …, *X*_50_
2. Simulated coefficients for confounders between (0,1) *α*_0_, …, *α*_50_
3. Simulated 5 instrumental variables that affect treatment only *X*_51_, …, *X*_55_ and the corresponding coefficients *α*_51_, …, *α*_55_
4. Generated treatment *T* by

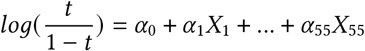

5 Set treatment effect coefficient *α*_*t*_
6 Simulated one risk factor that affect outcome only *X*_56_
7 Simulated coefficients *β*_0_, …, *β*_50_, *β*_56_ for outcome
8 Generated outcome *Y* by

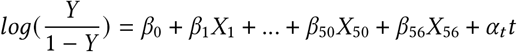

##### Nonlinear data

1. Simulated 25 confounders *X*_1_, …, *X*_25_ from normal distributions, where *X*_1_, …*X*_5_ were simulated from multivariate normal distribution and *X*_6_, …*X*_25_ were simulated from standard normal distribution.
2. Took cubic term of first five confounders, 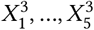 accounting for nonlinearity
3. Simulate 25 confounders from *X*_26_, …, *X*_50_ from binomial distributions, where *X*_26_, …, *X*_30_ were simulated from multivariate binomial distribution and *X*_31_, …*X*_50_ were simulated from binomial distribution inde-pendently
4. Generated 5 interactions between normal distribution confounders and binomial distribution confounders *X*_51_, …, *X*_55_ :

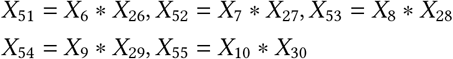

5 Generated 5 interaction terms between two normal distribution confounders *X*_56_, …, *X*_60_:

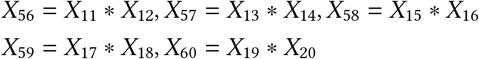

6 Generated 5 interaction terms between cubic terms and binomial distribution confounders *X*_61_, …, *X*_65_:

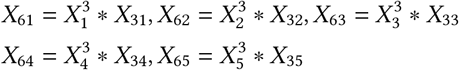

7 Simulated coefficients for confounders between (0,1) *α*_0_, …, *α*_65_
8 Simulated 5 instrumental variables that affect treatment only *X*_66_, …, *X*_70_ and the corresponding coefficients *α*_66_, …, *α*_70_
9 Generated treatment *T* by

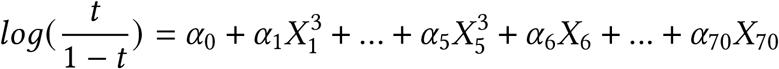

10 Set treatment effect coefficient *α*_*t*_
11 Simulated one risk factor that affect outcome only *X*_71_
12 Simulated coefficients *β*_0_, …, *β*_65_, *β*_71_ for outcome
13 Generated outcome *Y* by

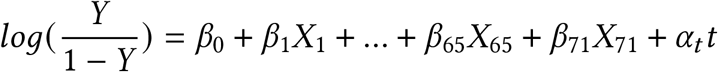

## REFERENCES

[1] Austin PC. The Relative Ability of Different Propensity Score Methods to Balance Measured Covariates Between Treated and Untreated Subjects in Observational Studies. Medical decision making. 2009;29(6):661–77.

[2] Austin PC. An Introduction to Propensity Score Methods for Reducing the Effects of Confounding in Observational Studies. Multivariate behavioral research. 2011;46(3):399–424.

[3] Ali MS, Groenwold RH, Klungel OH. Best (but oft-forgotten) practices : propensity score methods in clinical nutrition research. The American journal of clinical nutrition. 2016;104(2):247–58.

[4] Hajage D, Tubach F, Steg PG, Bhatt DL, De Rycke Y. On the use of propensity scores in case of rare exposure. BMC medical research methodology. 2016;16(1):38–8.

[5] Arbogast PG, Ray WA. Performance of Disease Risk Scores, Propensity Scores, and Traditional Multivariable Outcome Regression in the Presence of Multiple Confounders. American Journal of Epidemiology. 2011 07;174(5):613–20. Available from: https://doi.org/10.1093/aje/kwr143.

[6] Xu S, Shetterly S, Cook AJ, Raebel MA, Goonesekera S, Shoaibi A, et al. Evaluation of propensity scores, disease risk scores, and regression in confounder adjustment for the safety of emerging treatment with group sequential monitoring. Pharmacoepidemiology and Drug Safety. 2016;25(4):453–61.

[7] Glynn RJ, Gagne JJ, Schneeweiss S. Role of disease risk scores in comparative effectiveness research with emerging therapies. Pharmacoepidemiology and drug safety. 2012;21(S2):138–47.

[8] Wyss R, Ellis AR, Brookhart MA, Jonsson Funk M, Girman CJ, Simpson RJ, et al. Matching on the disease risk score in comparative effectiveness research of new treatments. Pharmacoepidemiology and Drug Safety. 2015;24(9):951–61.

[9] Cannas M, Arpino B. A comparison of machine learning algorithms and covariate balance measures for propensity score matching and weighting. Biom J. 2019 07;61(4):1049–72.

[10] Westreich D, Lessler J, Funk MJ. Propensity score estimation: neural networks, support vector machines, decision trees (CART), and meta-classifiers as alternatives to logistic regression. J Clin Epidemiol. 2010 Aug;63(8):826–33.

[11] Lee BK, Lessler J, Stuart EA. Improving propensity score weighting using machine learning. Statistics in medicine. 2010;29(3):337–46.

[12] Greenland S. Invited commentary: variable selection versus shrinkage in the control of multiple confounders. American journal of epidemiology. 2008;167(5):523.

[13] Schneeweiss S, Rassen JA, Glynn RJ, Avorn J, Mogun H, Brookhart MA. High-dimensional Propensity Score Adjustment in Studies of Treatment Effects Using Health Care Claims Data. Epidemiology (Cambridge, Mass). 2009;20(4):512–22.

[14] Wang C, Wang S, Shi F, Wang Z. Robust Propensity Score Computation Method based on Machine Learning with Label-corrupted Data. 2018.

[15] Setoguchi S, Schneeweiss S, Brookhart MA, Glynn RJ, Cook EF. Evaluating uses of data mining techniques in propensity score estimation: a simulation study. Pharmacoepidemiology and Drug Safety. 2008;17(6):546–55.

[16] Pirracchio R, Resche-Rigon M, Chevret S. Evaluation of the propensity score methods for estimating marginal odds ratios in case of small sample size. BMC medical research methodology. 2012;12(1):70–0.

[17] Adelson JL, McCoach DB, Rogers HJ, Adelson JA, Sauer TM. Developing and Applying the Propensity Score to Make Causal Inferences: Variable Selection and Stratification. Frontiers in psychology. 2017;8:1413–3.

[18] Miettinen OS. STRATIFICATION BY A MULTIVARIATE CONFOUNDER SCORE. American journal of epidemiology. 1976;104(6):609–20.

[19] van Smeden M, de Groot J, Moons K, Collins GS, Altman DG, Eijkemans M, et al. No rationale for 1 variable per 10 events criterion for binary logistic regression analysis. BMC Medical Research Methodology. 2016;16.

[20] van der Laan MJ, Polley EC, Hubbard AE. Super Learner. Statistical Applications in Genetics and Molecular Biology. 2007;6(1). Available from: https://doi.org/10.2202/1544-6115.1309.

[21] Shortreed SM, Ertefaie A. Outcome-adaptive lasso: Variable selection for causal inference. Biometrics. 2017;73(4):1111–22.

[22] Benkeser D, Van Der Laan M. The Highly Adaptive Lasso Estimator. In: 2016 IEEE International Conference on Data Science and Advanced Analytics (DSAA); 2016. p. 689–96.

